# Real World Experience and Clinical Utility of EsoGuard® - Interim Data from the Lucid Registry

**DOI:** 10.1101/2023.09.26.23296162

**Authors:** Richard Englehardt, Jason B. Samarasena, Nikolai A. Bildzukewicz, Rachelle Hamblin, Victoria T. Lee, Suman Verma, Brian J. deGuzman, Lishan Aklog

## Abstract

**Background:** Barrett’s Esophagus (BE) is the only known precursor to esophageal adenocarcinoma (EAC), and guidelines exist for screening, surveillance, and treatment. However, historically most high-risk individuals have not been reliably screened, likely due to a combination of factors associated with patient/physician awareness and use of upper endoscopy (UE) as the traditional screening test. EsoGuard® (EG) is a DNA biomarker assay, and EsoCheck® (EC) is a non-invasive, swallowable capsule device designed to collect cells from a targeted region of the esophagus. EG and EC in combination offers a well-tolerated, accessible, in-office triage test to improve detection of BE in patients with multiple risk factors. The Lucid Registry captures real-world data from the commercial use of EC with EG, and we present an interim review of clinical utility data from the first 517 enrolled subjects.

**Methods:** Multicenter, prospective, registry designed to capture data from patients undergoing EC cell collection and EG testing in the commercial setting. Data collection consists of demographics, risk factors, test results, provider management, and early clinical outcomes (through a maximum of four months post-EG). This data snapshot includes subjects enrolled from the start of the registry (April 14, 2023), through August 16, 2023. The primary assessment of clinical utility was agreement between EG assay results and physicians’ decision on whether to refer the patient for subsequent UE. The relationship between BE/EAC risk factors and EG positivity rates was assessed.

**Results:** Among 517 subjects enrolled, average age was 47.9±14.3 years, 47.2% had history of gastroesophageal reflux disease (GERD), and 63.8% had a minimum of 3 established BE risk factors (i.e., met American Gastroenterological Association (AGA) criteria for screening). 58.8% of subjects were firefighters; when firefighting i.e., occupational exposure to smoke and carcinogens is treated as an additional BE/EAC risk factor (+) those of the AGA, 81.2% of the study population had ≥3 risks, making up the “AGA(+)” cohort. EG positivity was 14.1%. 437 subjects contributed data for the clinical utility endpoint: agreement between positive EG results and subsequent referral for UE was 100%; agreement between negative EG results and non-referral for UE was 99.4%; concordance between EG results and UE referral decisions was 97.9%. These findings were comparable between the AGA and AGA(+) cohorts.

**Conclusions:** Experience from the Lucid Registry demonstrates that physicians who have adopted EC/EG in the commercial setting are reliably utilizing EG as a triage test to inform decision making on which patients to refer for further endoscopic evaluation of BE.

## Introduction

Esophageal adenocarcinoma (EAC) is a highly lethal cancer and for whom most patients present at late stages of disease.[1] It is the most common cancer of the esophagus in the United States, with climbing numbers in the past several decades, particularly in white males, for whom incidence has increased nearly 6-fold.[2-4] National statistics estimate there will be 21,560 new cases of esophageal cancer in 2023, resulting in approximately 16,120 deaths.[5] The 5-year relative survival is estimated at 21.7%, despite treatments including surgical resection, chemotherapy, and radiation therapy. Barrett’s Esophagus (BE) is the only known direct precursor to EAC and has well defined risk factors that characterize a “high-risk” population to develop BE.[6] In contrast to EAC, BE can be successfully treated using minimally invasive approaches such as radiofrequency or cryotherapy ablation with 80-90% success rates, highlighting the importance of early diagnosis.[7, 8]

Despite this, literature shows less than 20% of patients in the U.S who are diagnosed with EAC have a preceding diagnosis of BE, suggesting that current screening strategies are woefully inadequate.[9] This clinical gap could be due to poor understanding from referring providers around the disease, risk factors, and its association with malignancy, and/or patient reluctance to undergo traditional screening upper endoscopy (UE), which many perceive as uncomfortable and invasive. Further, there are limitations to endoscopy with forceps biopsy as the initial approach to screening. Aside from the low rates of referral to gastroenterologists for screening UE, biopsy may miss up to 50% of BE cases due to a combination of low adherence to structured biopsy protocols, sampling error, and other factors.[10] As such, more sensitive and easier to access methods of BE detection are needed.

EsoCheck® (EC) is a non-endoscopic, swallowable, balloon-based capsule device that allows for circumferential esophageal mucosal cell sampling, and when paired with the EsoGuard® (EG) biomarker assay, offers a minimally invasive strategy recognized by both the American College of Gastroenterology (ACG) and American Gastroenterological Association (AGA) as a reasonable alternative to UE for BE screening.[6, 11] EG, when used to analyze samples collected using EC, is not intended as a replacement for UE to assess ***known*** esophageal pathology. It may, however, be beneficial as a quick, easy to implement, and well-tolerated triage test that can be used in both a primary care and specialty setting to assist in the decision-making process for patients deemed at increased risk of BE. The goal of the ongoing, Lucid Diagnostics (EsoGuard) Registry (sponsored by Lucid Diagnostics Inc., New York, NY) is to collect real-world data from commercial EG experience to evaluate patient experience and satisfaction, and the impact of test results on health care provider’s management decisions. All patients undergoing EG testing in the commercial setting whose cell samples were collected using EC by Lucid personnel and consented to contributing data for the Registry were included. The data presented here is for the first 517 subjects enrolled into the study, among whom 437 had sufficient information from which the clinical utility of EG can be gleaned based on assay results and subsequent physician management decisions.

## Methods

To evaluate the utility of EG as a tool in the diagnosis of BE, patient demographics, risk factors, EG results, and provider management decisions were recorded and analyzed in a prospective, observational registry. The tolerability of EC was also assessed by documenting the severity of patient gag response and the number of failed cell collections. Any patients for whom his/her physician made an independent clinical decision to screen for BE using EC/EG were invited to participate in the Lucid Diagnostics Registry. Subjects were recruited from Lucid Test Centers (LTCs), satellite testing locations, and physician, community, or employer organized health fairs/screening events from April 14^th^, to August 16^th^, 2023.

The study was conducted according to the guidelines of the Declaration of Helsinki and approved by the WCG Institutional Review Board (IRB tracking number 20226705). All participating individuals signed informed consent prior to EC and collection of any study information. Enrollment is ongoing and the trial is registered on ***clinicaltrials***.***gov*** as NCT05965999.

### EsoCheck® and EsoGuard® (EC/EG)

EsoCheck® is an FDA 510K cleared, non-endoscopic, swallowable device designed for the circumferential collection and retrieval of surface cells from the esophagus (**Figure 1**). The unique, balloon-capsule technology allows for easy swallowing, non-traumatic targeted cell sampling, and protection of the cell sample during retrieval of the device through the upper esophagus and oropharynx. It is cleared for use in the general population of individuals 12 years of age or older. EsoGuard® is a laboratory developed test (LDT) performed in a Clinical Laboratory Improvement Amendment (CLIA) certified and College of American Pathologists (CAP) accredited lab that utilizes set of genetic assays and algorithms which examines the presence of cytosine methylation at 31 different genomic locations on the vimentin (VIM) and Cyclin-A1 (CCNA1) genes. EG has been clinically validated in a developmental study published in 2018 and shown to have a >90% sensitivity and >90% specificity in detection of BE or EAC.[12] EG results are reported in a binary fashion i.e., positive or negative, indicating presence or absence of sufficient methylation abnormalities to suggest diagnosis of BE or EAC. Infrequently, cell samples may have DNA “Quantity Not Sufficient” for EG analysis and are reported as “QNS,” or the samples may have quality issues prohibiting analysis which are reported as “Unevaluable” results. If this occurs, patients have the option of repeating testing with a new cell sample.

**Figure 1.**
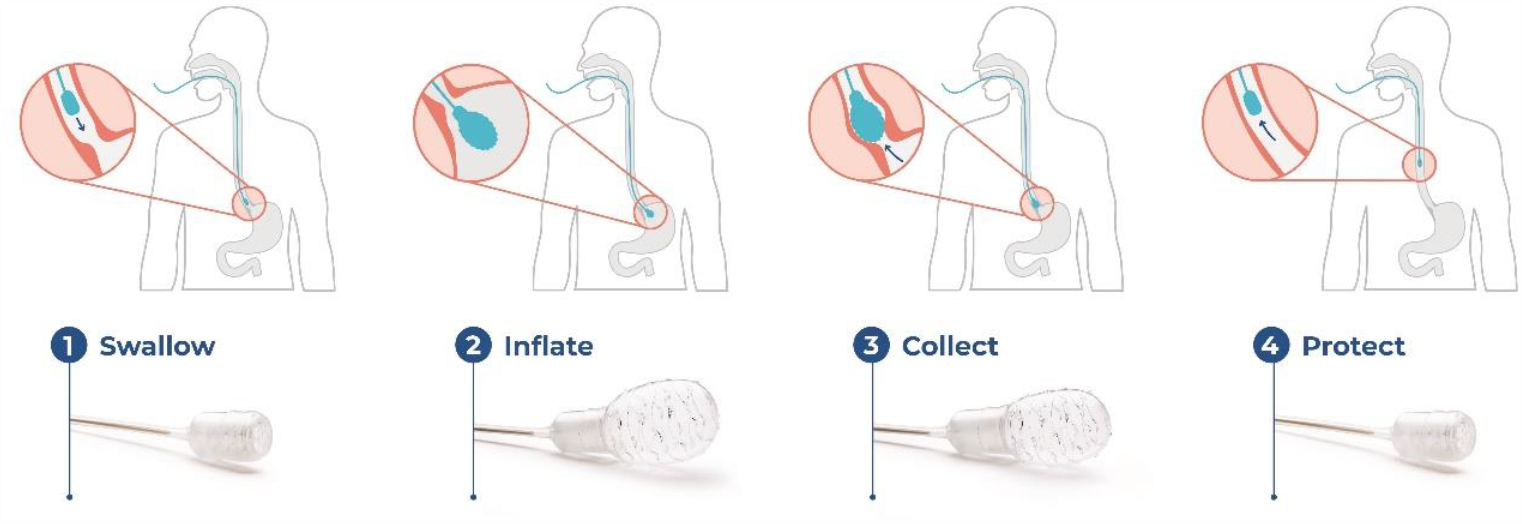
EsoCheck® Device and Cell Collection Process.

### Statistical Analysis

Subjects unable to swallow the EC device could not contribute cellular DNA for EG analysis; these subjects are included in the summary of enrollment demographics, but do not contribute to the clinical utility endpoint. Similarly, subjects without binary EG results (e.g., QNS or unevaluable) were included in overall data analysis but did not contribute to the primary clinical utility endpoints.

The primary analysis of clinical utility in this study was the positive agreement between EG positive (+) results and the decision to proceed with UE. Additional analyses included a) negative agreement between EG negative (-) results and the physician’s decision *not* to refer for UE, and b) overall **concordance** between the EG result and provider endoscopy decisions.

Positive agreement was calculated as the percentage of patients with EG(+) results who are referred for confirmatory UE; negative agreement was calculated as the percentage of patients with EG(-) results who are not referred for any UE.

Continuous variables were summarized using the number of observations (n), mean, standard deviation (SD), median, minimum, and maximum, along with total number of patients contributing values. Categorical variables were described by frequency of counts and percentages. The total number of applicable subjects (N) were used as the denominator for percent calculations unless stated otherwise within a table footnote. Binomial exact two-sided 95% confidence interval were calculated wherever relevant.

## Results

At the time of data snapshot, 517 subjects had signed informed consent, and distribution is provided in **Figure 2**. One subject was pending documentation of his/her EC cell collection details and aside from demographic and BE/EAC risk factor information, was excluded from data analysis. Only two individuals (2/516; 0.4%) were unable to tolerate the EC cell collection. Among the remaining 99.6% (514/516) who successfully completed EC to provide a cell sample for the Lab, clinical utility data was available for 437 (these subjects had both binary EG results and a physician decision on UE referral); these individuals contributed to the primary and secondary endpoint analyses.

**Figure 2.**
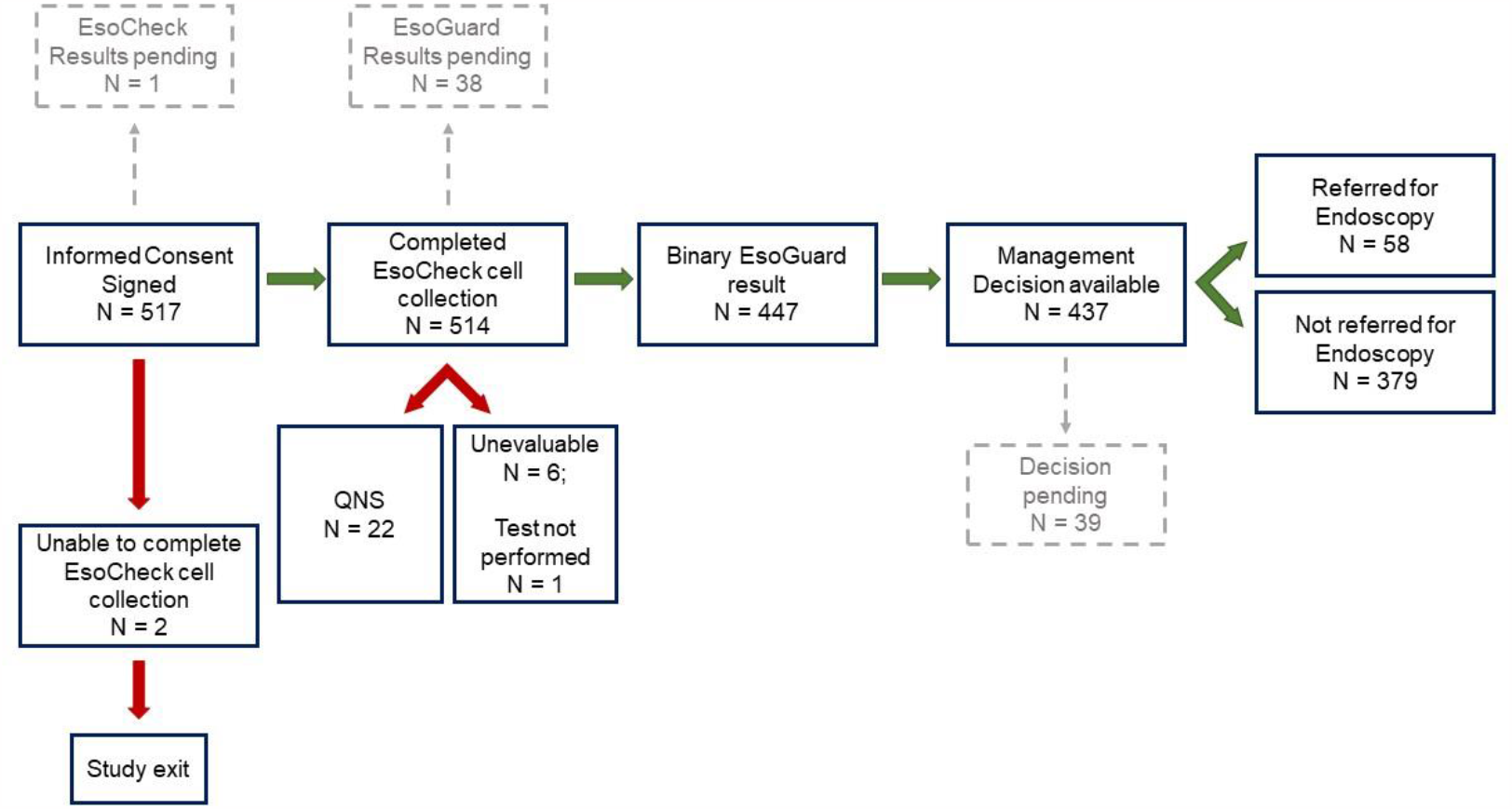
Subject Distribution.

An overview of demographics and BE risk factors is provided in **Table 1**. Not all 517 enrolled subjects had complete demographic/risk information at the time of this data snapshot, so analyses were performed on available datapoints. The mean age was 47.9 years (SD±14.3), 74.6% (385/516) were male, and 62.3% (322/517) were of White (Caucasian, non-Hispanic) race.

**Table 1.** Subject Baseline Characteristics.

| Characteristics | Overall<br>(N = 517*) |
| --- | --- |
| Age (at time of EsoCheck cell collection; years) |  |
| Mean ± SD | 47.9±14.3 (516) |
| Median (Q1, Q3) | 46.0 (37.0,58.0) |
| (Min, Max) | (20.0,88.0) |
| Biological Sex |  |
| Female | 25.4% (131/516) |
| Male | 74.6% (385/516) |
| White (Caucasian) Non-Hispanic |  |
| No | 37.7% (195/517) |
| Yes | 62.3% (322/517) |
| White (Caucasian) Hispanic |  |
| No | 69.8% (361/517) |
| Yes | 30.2% (156/517) |
| Black (or African American) |  |

**Table 1. Subject Baseline Characteristics**
| **Characteristics** | **Overall**<br>**(N = 517\*)** |
| --- | --- |
| No | 95.4% (493/517) |
| Yes | 4.6% (24/517) |
| American Indian or Alaskan Native |  |
| No | 99.8% (516/517) |
| Yes | 0.2% (1/517) |
| Asian (Asian Indian; Chinese; Filipino; Japanese; Korean; Vietnamese; Other Asian) |  |
| No | 97.3% (503/517) |
| Yes | 2.7% (14/517) |
| Native Hawaiian or other Pacific Islander |  |
| No | 99.4% (514/517) |
| Yes | 0.6% (3/517) |
| Cigarette Smoker (past or present)? |  |
| No | 67.5% (349/517) |
| Yes | 32.5% (168/517) |
| If positive history of cigarette smoking, indicate current status: |  |
| Current | 36.3% (61/168) |
| Former | 63.7% (107/168) |
| History of gastroesophageal reflux disease (GERD)? |  |
| No | 52.8% (273/517) |
| Yes | 47.2% (244/517) |
| If positive history of GERD, indicate number of years with disease |  |
| Mean ± SD | 9.8±8.9 (243) |
| Median (Q1, Q3) | 6.0 (4.0,12.0) |
| (Min, Max) | (0.0,50.0) |
| If positive history of GERD, how many times per week are symptoms experienced? |  |
| Mean ± SD | 3.1±2.2 (242) |
| Median (Q1, Q3) | 3.0 (1.0,5.0) |
| (Min, Max) | (0.0,7.0) |
| Are GERD symptoms controlled with the acid-controlling medications? |  |
| No | 20.5% (41/200) |
| Yes | 79.5% (159/200) |
| First degree family members with known diagnosis of Barrett's Esophagus (BE) and/or esophageal adenocarcinoma (EAC)? |  |
| No | 94.4% (487/516) |
| Yes | 5.6% (29/516) |
| History of occupational and/or environmental exposure to agents that may increase risk of BE or EAC? |  |

**Table 1. Subject Baseline Characteristics**
| Characteristics | Overall<br>(N = 517*) |
| --- | --- |
| No | 41.2% (213/517) |
| Yes | 58.8% (304/517) |
| Obese (defined as BMI $\geq 30\text{kg/m}^2$) | |
| No | 64.2% (332/517) |
| Yes | 35.8% (185/517) |
| Body mass index (BMI) |  |
| Mean ± SD | 29.2±5.2 (517) |
| Median (Q1, Q3) | 28.3 (25.8,31.7) |
| (Min, Max) | (16.6,50.1) |
| **AGA Cohort** (3 or more *established* risk factors as defined by National Societies) |  |
| No | 36.2% (187/517) |
| Yes | 63.8% (330/517) |
| **AGA (+) Cohort** (3 or more risk factors, including occupational/environmental exposure as a presumed EAC risk factor) |  |
| No | 18.8% (97/516) |
| Yes | 81.2% (419/516) |
| 4 or more *established* risk factors as defined by National Societies |  |
| No | 64.0% (331/517) |
| Yes | 36.0% (186/517) |
| 4 or more risk factors, including occupational/environmental exposure as a presumed EAC risk factor |  |
| No | 46.2% (239/517) |
| Yes | 53.8% (278/517) |
| Number of Risk Factors (any) |  |
| Mean ± SD | 3.6±1.3 (514) |
| Median (Q1, Q3) | 4.0 (3.0,4.0) |
| (Min, Max) | (0.0,8.0) |

Green highlighted boxes denote *established* risk factors as defined by National Societies; age is an established risk factor if >50 years
\*Total enrolled population N=517, among which all individuals contributed at least some demographic/risk factor information; however, not all subjects had complete information at the time of data snapshot thus the (n) in any individual row within this table may vary

Although <50% of subjects reported a history of gastroesophageal reflux disease (GERD), the average duration of symptoms in the GERD cohort was long, at nearly 10 years, and average symptom frequency was high, at approx. 3 times per week.

Patients with three (3) or more established risk factors for BE/EAC (as defined by the AGA and ACG in their screening guidelines) accounted for 63.8% (330/517) of the Registry population. The Registry also captured information on a special category of patients: 58.8% (304/517) of participants were firefighters, as denoted by a history of “*occupational/environmental exposure to carcinogenic chemicals*.” Most of these individuals were tested as part of department or community sponsored health fairs. When “*occupational/environmental exposure to carcinogenic chemicals*” was counted as a BE/EAC risk factor, 81.2% (419/516) of the study population had at least three (3) risk factors and met what our authors refer to as the “***AGA(+)***” criteria for BE/EAC testing (i.e., AGA-defined risk factors plus(+) occupational/environmental exposure as a potential risk).

### EsoCheck Cell Collection Procedure

One subject was pending documentation of his/her EC cell collection information at the time of data snapshot. All except two of the other enrolled subjects (514/516; 99.6%) successfully completed the EC cell collection (**Table 2**). The two (0.4%) subjects unable to swallow the device to provide cell samples for EG DNA analysis were exited from the study early.

**Table 2.** EsoCheck Cell Collection Characteristics.

| Characteristics | Overall<br>(N = 517*) |
| --- | --- |
| Mallampati Score[13] (device administrator assessment) |  |
| Score 1 | 29.1% (150/516) |
| Score 2 | 38.4% (198/516) |
| Score 3 | 24.0% (124/516) |
| Score 4 | 8.5% (44/516) |
| Was EsoCheck cell collection successful? |  |
| No | 0.4% (2/516) ↓ |
| Yes | 99.6% (514/516) |
| Time required for cell collection (*from start of device swallowing to time of complete device retrieval; seconds*) |  |
| Mean ± SD | 104.4±71.8 (514) |
| Median (Q1, Q3) | 60.0 (60.0,120.0) |
| (Min, Max) | (60.0,900.0) |
| Subject gag response (device administrator assessment; scale developed by and standardized among Lucid Clinical Services Staff) |  |
| Gag Score 1<br>(no or minimal gagging) | 36.6% (189/516) |
| Gag Score 2<br>(mild gagging) | 29.8% (154/516) |
| Gag Score 3<br>(moderate gagging) | 20.3% (105/516) |
| Gag Score 4<br>(severe gagging but able to complete cell collection) | 13.0% (67/516) |
| Gag Score 5<br>(severe gagging and unable to complete cell collection) | 0.2% (1/516)↓ |

\*For subjects who required more than one collection attempt, only the latest-most attempt was included in the count; one subject who was enrolled in the study (1/517) was pending documentation of his/her EsoCheck cell collection information at the time of data snapshot and thus excluded from the counts within this table
↓One subject who failed to swallow the EsoCheck device was documented with Gag response score <5, as the gagging was not visually assessed as severe, however the subject experienced sudden vomiting and the device was thus retrieved without adequate cell sample

The mean cell collection time was 104 seconds (1.73min), SD±71.8 seconds; the median cell collection time was 60 seconds (1 min). Most subjects had a Mallampati score of 3 or less.

The ‘Gag response score’ was a 5-point scale utilized by the device administrator to assess patient tolerability, with a score of 1 indicating the best tolerability (no gagging or minimal gagging) and a score of 5 indicating the worst tolerability (such severe gagging that cell collection could not be completed). Most patients (448/516; 86.8%) tolerated the cell collection well, with a gag response score of 3 or less.

### EsoGuard Results and Clinical Utility Evaluation

Of the 517 enrolled subjects, 476 had their EG results documented in the Registry database at the time of data snapshot (**Table 3**). Of note, more EG results may have been reported by the Central Lab to the ordering provider(s) but delayed data entry into the Registry database accounts for some discrepancies between the number of subjects in **Table 1** compared to **Table 3**. The EG positivity rate was 14.1% (67/476), and 79.8% (380/476) of patients were EG negative, 4.6% (22/476) of subjects who had insufficient DNA quantity in their cell samples for analysis (QNS), and 1.3% (6/476) of samples were Unevaluable due to quality failure. One sample (0.2%; 1/476) was not yet analyzed due to administrative or other sample issues. This resulted in 93.9% (447/476) of samples with binary EG results.

**Table 3.**
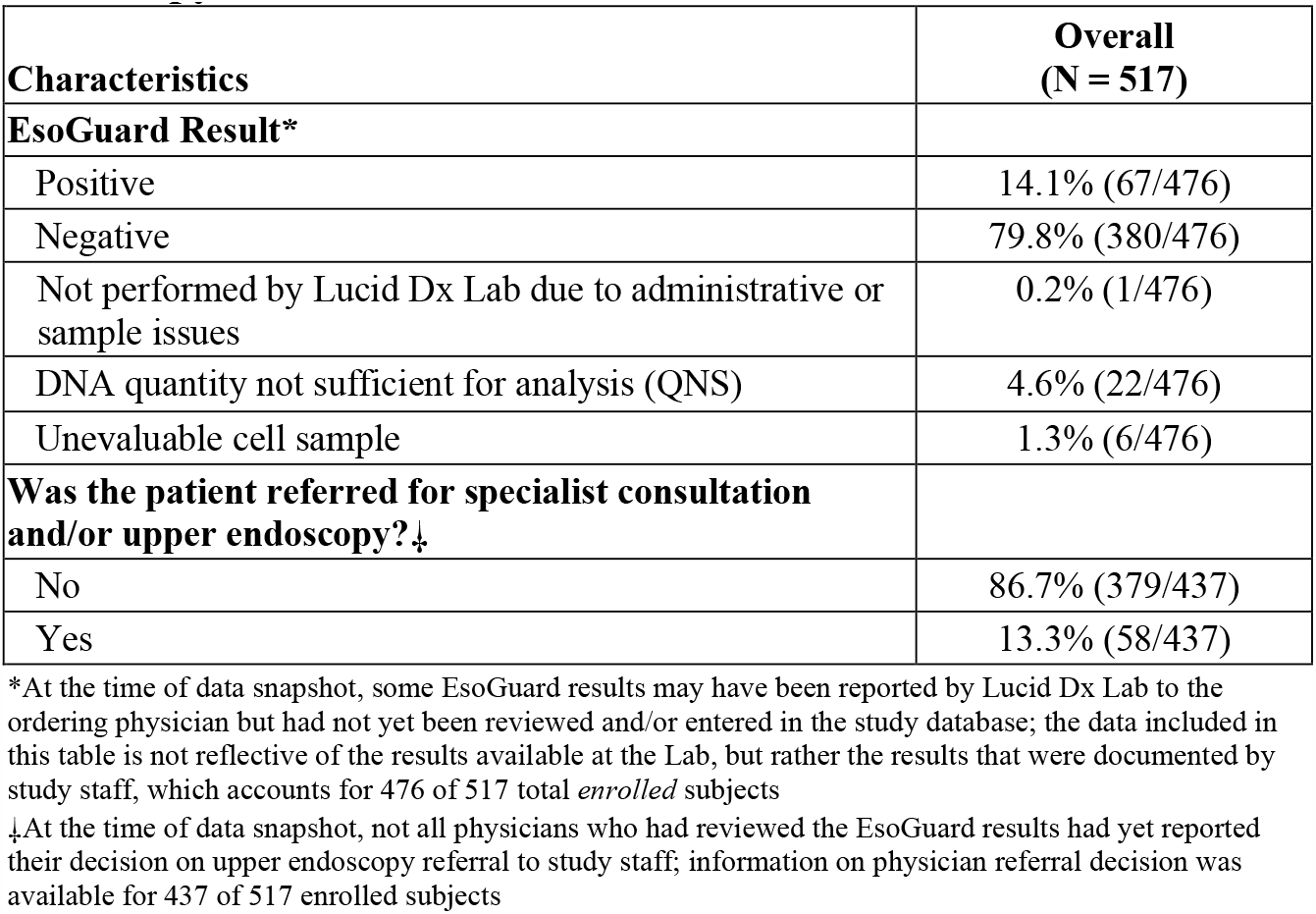
EsoGuard Result & Physician Decisions on Upper.

| **Characteristics** | **Overall**<br>**(N = 517)** |
| --- | --- |
| **EsoGuard Result\*** |  |
| Positive | 14.1% (67/476) |
| Negative | 79.8% (380/476) |
| Not performed by Lucid Dx Lab due to administrative or sample issues | 0.2% (1/476) |
| DNA quantity not sufficient for analysis (QNS) | 4.6% (22/476) |
| Unevaluable cell sample | 1.3% (6/476) |
| **Was the patient referred for specialist consultation and/or upper endoscopy?‡** |  |
| No | 86.7% (379/437) |
| Yes | 13.3% (58/437) |

\*At the time of data snapshot, some EsoGuard results may have been reported by Lucid Dx Lab to the ordering physician but had not yet been reviewed and/or entered in the study database; the data included in this table is not reflective of the results available at the Lab, but rather the results that were documented by study staff, which accounts for 476 of 517 total *enrolled* subjects
‡At the time of data snapshot, not all physicians who had reviewed the EsoGuard results had yet reported their decision on upper endoscopy referral to study staff; information on physician referral decision was available for 437 of 517 enrolled subjects

Referral decisions based on subject risk cohort: AGA vs. AGA(+) are summarized in **Table 4**. A decision from the ordering physician about specialist/UE referral was available for 437 subjects. In addition to the one subject for whom the EG assay was not performed due to administrative/other sample issues, there were two subjects with unevaluable samples, and 14 subjects with QNS results who did not have a decision on UE referral, likely due to anticipation of EG re-test. Among the subjects with binary EG results, 409 (55 positive, 354 negative) had physician decisions on specialist/UE referral.

**Table 4.** EsoGuard Results and Decisions on Endoscopy Referral by Risk Cohort.

| Physician Decision on UE Referral | Overall |  | Positive |  | Negative |  | QNS |  | Unevaluable |  |
| --- | --- | --- | --- | --- | --- | --- | --- | --- | --- | --- |
|  | % (n/N) | 95% CI | % (n/N) | 95% CI | % (n/N) | 95% CI | % (n/N) | 95% CI | % (n/N) | 95% CI |
| Full Study Cohort<br>(n = 475) |  |  |  |  |  |  |  |  |  |  |
| Not referred | 86.7%<br>(378/436) | [83.1%,89.7%] | 0.0%<br>(0/55) | [0.0%,6.5%] | 99.4%<br>(352/354) | [98.0%,99.9%] | 100.0%<br>(21/21) | [83.9%,100.0%] | 83.3%<br>(5/6) | [35.9%,99.6%] |
| Referred | 13.3%<br>(58/436) | [10.3%,16.9%] | 100.0%<br>(55/55) | [93.5%,100.0%] | 0.6%<br>(2/354) | [0.1%,2.0%] | 0.0%<br>(0/21) | [0.0%,16.1%] | 16.7%<br>(1/6) | [0.4%,64.1%] |
| Cohort meeting AGA screening criteria::<br>(n = 294*) |  |  |  |  |  |  |  |  |  |  |
| Not referred | 84.1%<br>(217/258) | [79.1%,88.3%] | 0.0%<br>(0/38) | [0.0%,9.3%] | 99.0%<br>(202/204) | [96.5%,99.9%] | 100.0%<br>(14/14) | [76.8%,100.0%] | 50.0%<br>(1/2) | [1.3%,98.7%] |
| Referred | 15.9%<br>(41/258) | [11.7%,20.9%] | 100.0%<br>(38/38) | [90.7%,100.0%] | 1.0%<br>(2/204) | [0.1%,3.5%] | 0.0%<br>(0/14) | [0.0%,23.2%] | 50.0%<br>(1/2) | [1.3%,98.7%] |
| Cohort meeting AGA(+) screening criteria::<br>(n = 377) |  |  |  |  |  |  |  |  |  |  |
| Not referred | 84.9%<br>(287/338) | [80.6%,88.6%] | 0.0%<br>(0/48) | [0.0%,7.4%] | 99.3%<br>(269/271) | [97.4%,99.9%] | 100.0%<br>(15/15) | [78.2%,100.0%] | 75.0%<br>(3/4) | [19.4%,99.4%] |
| Referred | 15.1%<br>(51/338) | [11.4%,19.4%] | 100.0%<br>(48/48) | [92.6%,100.0%] | 0.7%<br>(2/271) | [0.1%,2.6%] | 0.0%<br>(0/15) | [0.0%,21.8%] | 25.0%<br>(1/4) | [0.6%,80.6%] |

\*36 out of 330 subjects within this risk cohort were pending UE referral decisions at the time of data snapshot
‡42 out of 419 subjects within this risk cohort were pending UE referral decisions at the time of data snapshot
::Some, but not all subjects in the AGA(+) risk cohort also fell within the AGA risk cohort, whereas all subjects in the AGA risk cohort also met criteria for the AGA(+) cohort

All EG positive patients were referred for UE, irrespective of whether patients were in the AGA risk cohort vs. ‘AGA(+)’ cohort. Only two subjects with a negative EG result were referred for UE – all others did not proceed with further diagnostic work-up. These two subjects both met the AGA criteria for BE screening.

The primary clinical utility endpoint was calculated for subjects who had both a binary EG result and a physician decision on UE referral (**Table 5**). The ***positive agreement*** between an EG(+) test result and decision to refer the subject to specialist and/or UE was calculated at 100%, which was consistent across all cohorts. The ***Negative agreement*** for the full study population was 99.4%, which was nearly identical for the AGA and AGA(+) cohorts at 99.0% and 99.3% respectively. For subjects with ≥4 BE/EAC risk factors, the negative agreement was >98%. O***verall concordance*** between EG result and physician decision on UE referral was 97.9%.

**Table 5.** Primary Clinical Utility Endpoint(s) by Risk Category.

| Analysis Set | Subjects with Binary EG Result and a physician decision on UE referral (N) | Positive Agreement*<br>(95% CI) | Negative Agreement‡<br>(95% CI) | Concordance (95% CI)‡ |
| --- | --- | --- | --- | --- |
| Overall | 409 | 100.0%<br>(93.5%,100.0%) | 99.4%<br>(98.0%,99.9%) | 97.9%<br>(95.1%,100.0%) |
| AGA Cohort | 242 | 100.0%<br>(90.7%,100.0%) | 99.0%<br>(96.5%,99.9%) | 96.9%<br>(92.7%,100.0%) |

**Table 5. Primary Clinical Utility Endpoint(s) by Risk Category**
| Analysis Set | Subjects with Binary EG Result and a physician decision on UE referral (N) | Positive Agreement* (95% CI) | Negative Agreement‡ (95% CI) | Concordance (95% CI)‡ |
| --- | --- | --- | --- | --- |
| AGA(+) Cohort | 319 | 100.0%<br>(92.6%,100.0%) | 99.3%<br>(97.4%,99.9%) | 97.6%<br>(94.3%,100.0%) |
| Cohort with 4 or more *established* risk factors (*excludes occupational/environmental exposure as a risk*) | 139 | 100.0%<br>(88.8%,100.0%) | 98.1%<br>(93.5%,99.8%) | 95.9%<br>(90.4%,100.0%) |
| Cohort with $\geq 4$ risk factors for BE *including occupational/ environmental exposure* | 207 | 100.0%<br>(90.3%,100.0%) | 98.8%<br>(95.8%,99.9%) | 96.7%<br>(92.2%,100.0%) |

\*Primary Clinical Utility Endpoint
‡Secondary Clinical Utility Endpoint

When comparing characteristics of EG(+) vs. EG(-) subjects, there was a statistically significant difference in age and number of BE/EAC risk factors (**Table 6**). In general, the EG positive subjects were older (mean age of 57.8 years) and had more risk factors (average of 4.1) compared to EG negative subjects (mean age of 45.8 years and average 3.5 risk factors). Of similar statistical significance was the finding that EG positive patients were more likely than EG negative subjects to have ≥4 *established* BE risk factors.

**Table 6.** Comparison of Characteristics Between EsoGuard Positive vs. Negative Subjects.

| Characteristics | Overall<br>(N = 475) | EG Positive<br>(N = 67) | EG Non-Positive*<br>(N = 408) | p-Value |
| --- | --- | --- | --- | --- |
| Age (years) |  |  |  |  |
| Mean ± SD | 47.5±14.2 (474) | 57.8±14.1 (67) | 45.8±13.5 (407) | <.001 |
| Median (Q1, Q3) | 46.0 (36.0,57.0) | 56.0 (46.0,68.0) | 45.0 (35.0,54.0) |  |
| (Min, Max) | (20.0,88.0) | (22.0,88.0) | (20.0,81.0) |  |
| Number of Risk Factors (any, *including occupational/ environmental exposure*) |  |  |  |  |
| Mean ± SD | 3.6±1.3 (472) | 4.1±1.3 (67) | 3.5±1.3 (405) | <.001 |
| Median (Q1, Q3) | 4.0 (3.0,4.0) | 4.0 (3.0,5.0) | 3.0 (3.0,4.0) |  |
| (Min, Max) | (0.0,8.0) | (0.0,7.0) | (0.0,8.0) |  |
| **AGA Cohort** (3 or more established risk factors) N=330 |  |  |  | 0.021 |
| No | 38.1% (181/475) | 25.4% (17/67) | 40.2% (164/408) |  |
| Yes | 61.9% (294/475) | 74.6% (50/67) | 59.8% (244/408) |  |
| **AGA (+) Cohort** (3 or more risk factors; *includes occupational/environmental exposure as a risk*) N=419 |  |  |  | 0.028 |
| No | 20.5% (97/474) | 10.4% (7/67) | 22.1% (90/407) |  |
| Yes | 79.5% (377/474) | 89.6% (60/67) | 77.9% (317/407) |  |
| Cohort with 4 or more **established** risk factors (*excludes occupational/environmental exposure as a risk*) N = 186 |  |  |  | <.001 |
| No | 65.1% (309/475) | 46.3% (31/67) | 68.1% (278/408) |  |
| Yes | 34.9% (166/475) | 53.7% (36/67) | 31.9% (130/408) |  |
| Cohort with $\geq 4$ risk factors for BE *including occupational/ environmental exposure* N = 278 | | | | 0.009 |
| No | 47.6% (226/475) | 32.8% (22/67) | 50.0% (204/408) |  |
| Yes | 52.4% (249/475) | 67.2% (45/67) | 50.0% (204/408) |  |

\*Includes subjects who were either EG(-) or are pending EG results but have undergone successful EC cell collection and provided complete demographic/risk factor information

## Discussion

Among patients with GERD, the prevalence of BE is approximately 6%; this increases to over 12% in patients with GERD and at least one additional risk factor.[14] BE is the only known precursor to EAC, however current diagnostic approaches are only identifying approximately 20% of increased-risk patients. As a result, most cases of EAC are found in individuals without an established BE diagnosis, which means the window for early detection of the precancerous condition (BE) and intervention to prevent malignant progression was missed.[15-17] Despite multiple published societal guidelines endorsing BE screening, most patients who would benefit from testing are not being tested. There is evidence to suggest that patient factors and endoscopy access issues are contributory to some extent. Kolb et. al. utilized web-based surveys to collect information from GERD patients around how they perceive BE risk and the benefits of screening. About one fifth (20.4%) of patients admitted fear of discomfort as a barrier to undergoing screening UE, along with logistical considerations (e.g., scheduling, location, wait time, post-sedation/post-anesthesia needs etc.,); this concern was increased among patients who had never undergone a prior UE.[18] Interestingly, more Black respondents recognized the importance of screening and were more concerned about developing BE/EAC compared to White and other race groups, but more frequently reported difficulty with scheduling the procedure. This suggests that non-endoscopic approaches such as EsoCheck and EsoGuard (EC/EG) could reduce inequity and be instrumental in improving acceptability and accessibility of BE testing among individuals who might otherwise be unable or unwilling to comply with traditional diagnostic evaluation.

EG, when used to analyze esophageal cells collected with EC, was developed as a triage tool in the diagnosis of patients with BE; although data also demonstrates excellent ability to detect EAC,[12] the ultimate goal is early detection of BE (i.e., pre-malignant disease), which enables appropriate surveillance or treatment, and ultimately halting progression to malignancy. As a triage tool, EG would be performed as a first step in the diagnostic work-up, with the result informing the next step in patient management – namely UE. Within the population of the Lucid Registry, the EC cell collection process was highly efficient and successful, as demonstrated by the fact that only two of 516 subjects (0.4%) were unable to swallow the device and an average cell collection time of less than two minutes (104.4 seconds). Tolerability, as assessed by the patient’s ‘Gag response score’ was generally good, with most subjects (86.5%) evaluated to have a gag response score of 3 or less, and over a third having a score of 1 (no gag or minimal gag) (**Table 2**).

The target test population for EG is not a general screening population, as there are well-established criteria defining patients at increased risk for BE. Instead, EG is recommended for patients with multiple risk factors - consistent with society guidelines and practice recommendations. It is also not intended to replace UE in patients with alarming or “red flag” symptoms (e.g., dysphagia, refractory GERD symptoms, etc.) that would warrant UE for non-screening purposes. In 2022 the AGA published their *Clinical Practice Update on New Technology and Innovation for Surveillance and Screening of BE*, in which they recommended screening for individuals with at least three established risk factors and endorsed non-endoscopic cell collection paired with a biomarker test as a reasonable alternative to UE as an initial test.

Among Registry participants, 63.8% had characteristics aligned with AGA recommendations, and 81.2% of participants met what we refer to as “AGA(+)” criteria, meaning they had three or more risk factors for BE/EAC if occupation as a firefighter (with frequent environmental exposure to smoke and other carcinogenic compounds) were counted as a risk factor. As an occupation, firefighting is known to increase an individual’s likelihood of developing a multitude of malignancies including esophageal cancer and was designated a Group 1 carcinogen by the International Agency for Research on Cancer (IARC) in July of 2022.[19] While not included in current society guidelines as an *established risk factor* for BE/EAC, an increased incidence of esophageal cancer-related deaths in firefighters – most frequently EAC - is demonstrated in the literature and likely attributable to their ongoing exposure to smoke and other toxic agents.[20, 21] In a study of California firefighters, the odds ratio (OR) for development of EAC in those of White race was 1.84, and 1.85 for firefighters of any race.[22] This is comparable to the association between BE and central obesity, with an OR of 1.88 when adjusted for BMI, and OR of 1.98 when unadjusted for BMI.[23]

Regarding BE/EAC risk factors, when the EG(+) vs. EG(-) populations were compared, it was noted that in general, the EG positive subjects were older and had more risk factors compared to EG negative subjects. The EG positivity rate for the Lucid Registry population (14.1%) aligns well with incidence of BE from the literature in a multi-risk factor population, which ranges between 5-15%.[24] When we evaluated the AGA risk cohort and the “AGA(+)” cohorts separately, we saw that positivity rates were similar, at 15.1% (50/330) and 14.3% (60/419) respectively. Although data comparing EG results to endoscopy findings is outside the scope of this Clinical Utility discussion, the consistency between EG positivity rates and published disease prevalence is overall reassuring.

Clinical utility data collected in the Lucid Registry showed that 100% of patients with positive EG results were recommended for confirmatory UE, while only two EG negative subjects (2/354, 0.6%; **Table 4**) were referred for UE. The clinical utility of a triage test is derived from its ability to influence provider decision-making – namely on next steps in diagnostic evaluation or in patient management. The positive agreement between EG(+) results and UE referral of 100% demonstrates that ordering physicians take the findings of a positive biomarker test as actionable. Perhaps more importantly, the negative agreement of >99% between EG(-) results and decision ***not*** to refer for UE, reflects physician confidence in the ability of EG to appropriately capture any patients with disease. It suggests physicians are sufficiently reassured by a negative result to not waste further resources on additional diagnostic work-up. Overall, concordance between EG results and provider decision on UE referral is very high and demonstrates a consistent pattern of appropriate utilization of EG as a triage test in the real-world setting. Additionally, in the cohort of Registry patients with ≥3 established BE/EAC risk factors it can be postulated that a negative EG resulted in net cost savings by deferring expensive screening UE in patients who would otherwise have met Association guidelines to be tested in this a manner.

Focusing on provider decision-making as the sole measure of clinical utility may be perceived as a study limitation. Patient behavior i.e., compliance with recommended management (namely endoscopy) was not included within this data snapshot, but is information being captured within the registry database and will be presented in future publications. This outcome was excluded from current analysis due to paucity of data, which is attributable to timing of the snapshot and the frequently protracted follow-up duration required to obtain this information (reflective of long lead-times for most UE scheduling). As discussed previously, the delays and difficulties associated with scheduling UE are factors contributing to poor patient acceptance of this procedure as an initial BE screening test. Details about the individual EG ordering providers were not collected, nor included in this analysis. This may be considered an additional limitation, however, it can be assumed that ordering providers are diverse in respect to geography, specialty (consisting of primary care providers, gastroenterologists, foregut surgeons, and laryngoscopists), and consist of both academic and non-academic practices, as this is what’s seen in the overall Lucid Diagnostics commercial market. Indeed, most commercial clients are from non-academic institutions in non-urban areas, which is reflective of the overall mission of this technology to increase accessibility of BE testing outside regions/practices where UE is most readily available as a screening tool.

In short, preliminary review of real-world data collected from the Lucid Diagnostics Registry demonstrates that providers are reliably utilizing the test as a triage tool to guide determination of which patients to refer or not refer for endoscopic evaluation of BE. This approach could enable broader outreach and more consistent testing of increased-risk patients, while also focusing UE resources on those patients with the highest pre-procedure probability of disease.

## Conclusions

Experience from the Lucid Diagnostics (EsoGuard) Registry demonstrates that physicians who have adopted EC/EG in the commercial setting are reliably utilizing EG to inform decision making on which patients to refer for further endoscopic evaluation of BE. This strategy may be an effective method for ensuring that more high-risk individuals are undergoing diagnostic testing, while also tailoring UE resource utilization.

## Data Availability

All data produced in the present study are available upon reasonable request to the authors

## Notes

### Competing Interest Statement

Authors R.E., J.B.S., N.A.B, and R.H have no competing interests to declare. Authors V.T.L., S.V., B.J.D., and L.A. are members of the Executive team at Lucid Diagnostics Inc. and own stock and/or options in the company.

### Funding Statement

The study was funded by Lucid Diagnostics Inc.

### Author Declarations

The study was reviewed by the WCG IRB with initial certificate of action granting ethical approval of the study on 22-December-2022, under study tracking number 20226705. Continuing review frequency will be performed annually.

